# Social and territorial inequalities in late HIV diagnosis in Greater Paris, France: The ANRS-MIE COINCIDE Study

**DOI:** 10.1101/2025.11.25.25340657

**Authors:** A Rojas-Chaves, M Mary-Krause, C Delpierre, Y Yazdanpanah, P Chauvin, F Caby COINCIDE Study Group

## Abstract

**Background:** Late HIV diagnosis (LD) remains a major challenge in controlling the HIV epidemic, contributing to higher morbidity, mortality, and ongoing transmission. We aimed to estimate its prevalence and to identify associated socio-territorial inequalities in Greater Paris.

**Methods:** This observational study included PLWH diagnosed during 2014-2021 and treated in all HIV hospital centers across Greater Paris. Seven transmission groups were defined according to birth region, gender, and transmission mode. Neighborhood deprivation was assessed using the French Deprivation Index (FDEP) based on residence place. Factors associated with LD were identified using multilevel modeling.

**Findings:** Among 8,346 PLHW, 58% were born abroad, 43% were MSM, and 16% lived in precarious housing (3% among French-born MSM (FMSM) vs. 26% and 30% among foreign-born heterosexual men (fHM) and women (fW)) Overall, 47% lived in the most deprived neighborhoods, reaching 64% among fHM and 62% among fW. Nearly half had LD (47%), ranging from 30% among FMSM, to 65%/57% among fHM/fW. In multivariate analysis, LD was associated with living in deprived neighborhoods (aOR=1.39, 95%CI [1.17-1.65]). After adjustment for transmission group, all groups showed higher LD risk than FMSM, and previous associations disappeared.

**Conclusion:** Living in deprived areas was associated with higher LD risk, driven by the concentration of high-risk populations (fHM and fW) and reflecting the socio-spatial segregation of PLWH. These findings highlight the need for innovative, geography-based, screening strategies that go beyond traditional risk-group approaches. Moreover, the seven-category “transmission group” classification represents a valuable social indicator for studying health inequalities among PLWH.

## Introduction

Late HIV diagnosis (LD) remains frequent and still represents a major public health challenge. In 2023, over half^1^ of people living with HIV (PLWH) in the European Union and European Economic Area (EU/EEA) had a CD4 count≤350/mm^3^ at diagnosis, and more than one third had a count ≤200/mm³. Despite advances in antiretroviral therapy (ART), low CD4 counts at diagnosis remain associated with higher mortality^2,3^, even after immune recovery^4^, and increased healthcare costs^5,6^ for several years after ART introduction. Beyond its clinical impact, LD fuels HIV epidemic, as undiagnosed and untreated people are more likely to transmit the virus. Early testing and prompt ART initiation allow reducing transmission risk to zero and are essential to ending the epidemic^7,8, 9^.

In Europe, LD disproportionately affects women, people over 40, and migrants from Southeast Asia and Sub-Saharan Africa^10,11^. While US studies associate LD to racial segregation, neighborhood cohesion, and socioeconomic deprivation^12,13,14^, European data are scarce and inconsistent. A French study found no association between socio-territorial deprivation and LD^15^, whereas others reported such an association in Switzerland and Germany^16,17^. Clarifying the influence of socio-territorial deprivation on HIV diagnosis is therefore crucial for guiding public health policies and improving access to care.

Greater Paris, characterized by marked socioeconomic disparities^18^, remains the most affected region in mainland France, accounting for 42% of new LD diagnoses in 2023 while hosting 19% of the population^19,20,21^.

The ANRS-MIE COINCIDE study has been designed to map new HIV diagnoses at a fine geographical scale in Greater Paris^21^, to estimate LD prevalence, and to identify associated socio-territorial inequalities. Here we specifically studied the relationship between socio-territorial deprivation and LD.

## Methods

### Study design and population

This observational study included all adults newly diagnosed with HIV-1 between 2014 and 2021, residing and receiving care in Greater Paris. Foreign-born PLWH with a plasma viral load<500copies/mL (suggesting prior diagnosis/treatment), and those with missing data were excluded (Figure 1).

**Figure 1.**
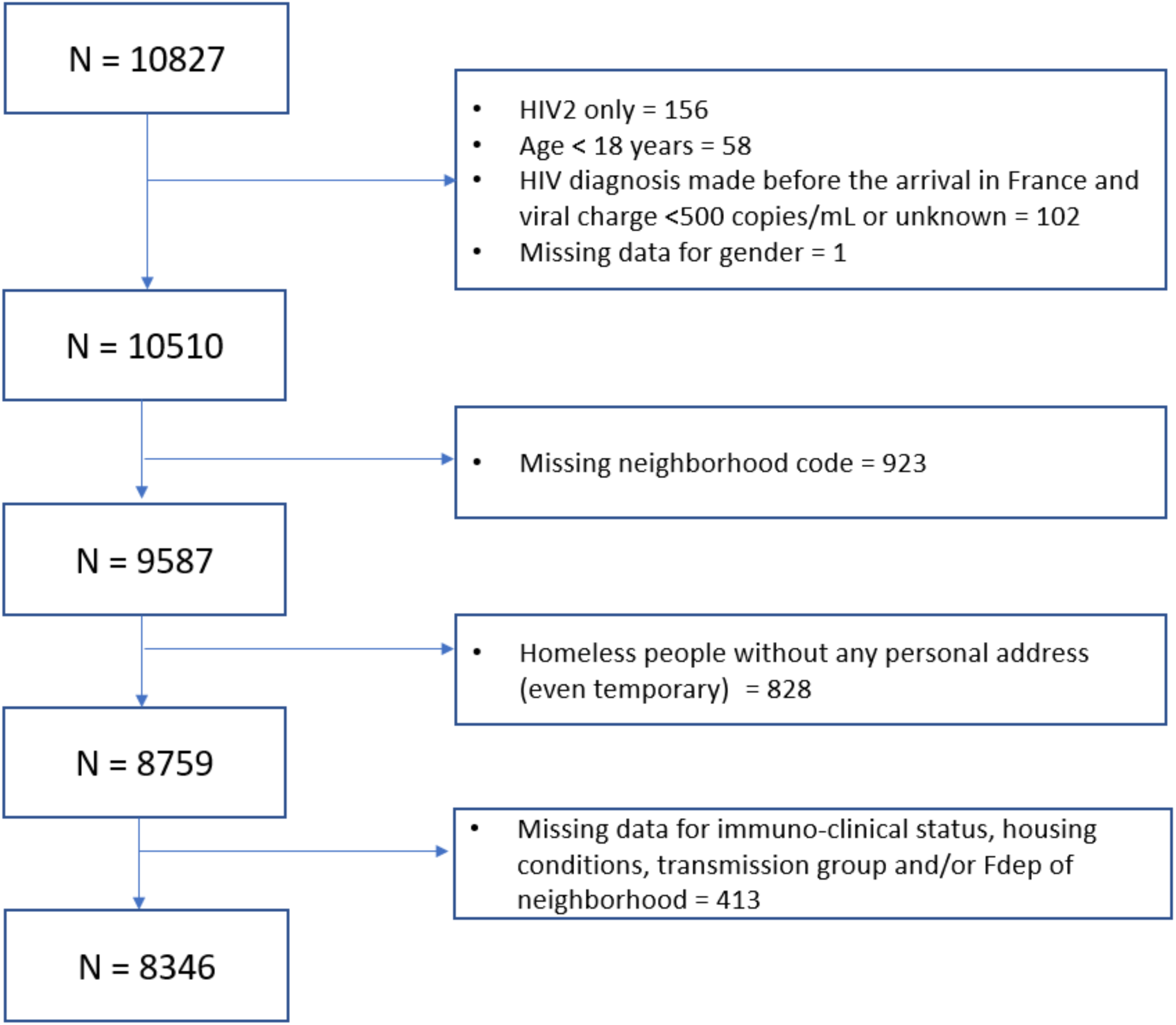
Flow-chart of the study population

### Data

Data were collected from the 61 HIV centers in Greater Paris, as part of the COINCIDE study (*CartOgraphies INfra-départementales des nouveaux diagnostiCs d’infection à VIH en Ile-DE-France*), supported by the ANRS-MIE (*Agence Nationale de Recherche sur le SIDA, les hépatites virales et les Maladies Infectieuses* Émergentes). These centers maintain prospective databases of all PLWH who provide written consent for sociodemographic, clinical, and biological data collection during routine care.

### Main outcome

The consensus definition of LD was applied throughout the study i.e., “having a clinical AIDS-defining event or CD4<350/mm³ at diagnosis, excluding primary infection***”*** ^10,22^.

### Main exposure

**Territorial variables** at the place of residence were provided by the French National Institute of Statistics and Economic Studies (INSEE), the National Health Data System, and the Regional Health Authority (ORS-IDF).

**Neighborhoods** corresponded to IRIS units^23^ (the smallest French census areas, with 2,500 inhabitants in 0.31 km^2^ on average). Socioeconomic deprivation was measured using the latest version (2015) French Deprivation Index (FDEP), combining four ecological variables^23^: unemployment rate, proportion of manual workers, proportion of high school graduates, and household median income. Each individual was assigned to an FDEP quintile (Q5=most deprived), weighted by the neighborhoods population in 2017.

**At the city level** (boroughs for inner Paris), three healthcare indicators were assessed as potential confounders of the FDEP-LD association: i/Accessibility to primary care (potential visits to general practitioner/inhabitant/year), ii/Presence of any municipal sexual health center, iii/ HIV screening rate (individuals with ≥1 HIV-test in 2021/1,000 inhabitants).

Territorial variables were assigned to each PLWH using geocoded residential address.

### Covariables

**Individual variables** included: year of diagnosis, gender, region of birth, transmission mode (heterosexual/men who have sex with men (MSM)/other), and at diagnosis: age, plasma HIV viral load, CD4 count, AIDS status, housing status (personal residence/precarious housing-homeless or temporarily housed by a third party), and residential zone (inner Paris, close suburbs, remote suburb).

Due to strong interactions between gender, transmission mode and country of birth, a composite “transmission group” variable was created, including seven categories: French-born MSM (FMSM), foreign-born MSM (fMSM), French-born heterosexual men (FHM), foreign-born heterosexual men (fHM), French-born women (FW), foreign-born women (fW), and transgender individuals.

### Statistical analysis

Categorical variables were expressed as n(%), and continuous variables as median(interquartile range-IQR). Group comparisons used the Chi-square test for proportions and the Kruskal-Wallis test for medians. Factors associated with LD were first tested using univariate logistic regression; those with p<0.20 were included in a multilevel model considering three levels: individual, neighborhood, and city. Seven sequential models (all adjusted for age) were built: **Model 1** included the FDEP score alone (neighborhood level); **Models 2-4** adding the three healthcare indicators (city level); **Models 5-7** adding individual factors (residential zone, precarious housing, transmission group). Fixed effects were expressed as adjusted Odds (aOR) ratios with 95% confidence intervals ([95% CI]), and random effects by intra-cluster correlation and conditional R².

As we hypothesized that both the FDEP and the transmission group were associated with LD, an additional analysis was carried out by dichotomizing the “transmission group” variable and comparing each category (except for transgender people due to small numbers) to the rest of the population. This analysis improved statistical power and aimed to clarify the FDEP-LD association while accounting for differences across transmission group categories.

### Cartographic representations

To visualize the FDEP-LD association, we mapped the proportion of LD in Paris inner-city, close and outer suburbs at the city level, overlaying them on the FDEP map.

All analyses and mapping were performed using RStudio (4.2.1).

## Results

### Study population

Between 2014 and 2021, 10,827 PLWH were newly diagnosed and initiated ART in Greater Paris. Of these, 2,481 were excluded (Figure 1). Excluded individuals were younger (median age 34), more often diagnosed in 2020-2021 (31%), less frequently French-born (22%), mostly from sub-Saharan Africa (56%), more often heterosexual (57%), in precarious housing (45%), and more frequently LD (52%). The final study population included 8,346 PLWH.

Tables 1-2 summarize individual and territorial characteristics overall and by transmission group. Overall, 58% were born-abroad (two thirds from sub-Saharan Africa). Among MSM, 49% lived in inner Paris, compared with 79% of foreign-born non-MSM in the suburbs. Precarious housing affected 16% of participants, more rarely French-born PLWH (3%-5%) than foreign-born PLWH, reaching 30% in fW. Nearly half (47%) resided in the most deprived neighborhoods (FDEP Q4-Q5) and was the most frequent in fHM (64%), fW (62%), and French-born women (55%). FMSM, fMSM and transgender participants more often lived in cities with better primary care access and higher HIV screening rates (64.0-64.8/1,000 inhabitants).

**Table 1.** Individual characteristics of the study population.

|  | Overall | French-born<br>MSM | Foreign-born<br>MSM | French-born<br>heterosexual<br>men | Foreign-born<br>heterosexual<br>men | French-born<br>women | Foreign-born<br>women | Transgender<br>people | *P* |
| --- | --- | --- | --- | --- | --- | --- | --- | --- | --- |
|  | N = 8346 | n = 2425 | n = 1136 | n = 696 | n = 1631 | n = 409 | n = 1981 | n = 68 |  |
| Year of diagnosis |  |  |  |  |  |  |  |  | <0.001 |
| 2014-2015 | 2705 (33%) | 825 (34%) | 313 (28%) | 212 (30%) | 529 (32%) | 131 (32%) | 689 (35%) | 11 (17%) |  |
| 2016-2017 | 2418 (29%) | 716 (30%) | 314 (28%) | 194 (28%) | 479 (30%) | 124 (30%) | 574 (29%) | 17 (25%) |  |
| 2018-2019 | 2189 (26%) | 614 (25%) | 330 (29%) | 197 (28%) | 426 (26%) | 102 (25%) | 495 (25%) | 25 (36%) |  |
| 2020-2021 | 1034 (12%) | 270 (11%) | 179 (15%) | 95 (14%) | 197 (12%) | 52 (13%) | 226 (11%) | 15 (22%) |  |
| Age (years) | 37 (29- 47) | 35 (27- 45) | 32 (27- 40) | 43 (32- 53) | 41 (34- 50) | 37 (27- 51) | 36 (30- 45) | 32 (27- 41) | <0.001 |
| Region of birth |  |  |  |  |  |  |  |  | <0.001 |
| France | 3545 (42%) | 2425 (100%) | - | 696 (100%) | - | 409 (100%) | - | 15 (22%) |  |
| Sub-Saharan Africa | 3234 (39%) | - | 298 (26%) | - | 1235 (76%) | - | 1692 (85%) | 9 (14%) |  |
| Middle East | 462 (6%) | - | 235 (21%) | - | 152 (9%) | - | 74 (4%) | 1 (1%) |  |
| Latin America | 277 (3%) | - | 191 (17%) | - | 28 (2%) | - | 23 (1%) | 35 (53%) |  |
| East Europe | 144 (2%) | - | 54 (5%) | - | 49 (3%) | - | 40 (2%) | 1 (1%) |  |
| Caribbean | 133 (1%) | - | 26 (2%) | - | 48 (3%) | - | 58 (3%) | 1 (1%) |  |
| Other | 550 (7%) | - | 332 (29%) | - | 119 (7%) | - | 94 (5%) | 5 (8%) |  |
| CD4 (/mm<sup>3</sup>) | 352<br>(185- 534) | 445<br>(289- 610) | 381<br>(243- 549) | 331<br>(130- 537) | 248<br>(101- 411) | 389<br>(201- 611) | 300<br>(153- 470) | 418<br>(287- 544) | <0.001 |
| Late HIV diagnosis<sup>1</sup> | 3902 (47%) | 724 (30%) | 451 (40%) | 340 (49%) | 1064 (65%) | 175 (43%) | 1127 (57%) | 24 (35%) | <0.001 |
| Advanced HIV diagnosis<sup>2</sup> | 2343 (28%) | 385 (16%) | 214 (19%) | 240 (34%) | 713 (44%) | 105 (26%) | 675 (34%) | 11 (16%) | <0.001 |
| Residential Zone |  |  |  |  |  |  |  |  | <0.001 |
| Paris inner-city | 2748 (33%) | 1171 (48%) | 558 (49%) | 160 (23%) | 340 (21%) | 73 (18%) | 415 (21%) | 31 (46%) |  |
| Close suburbs | 3244 (39%) | 763 (31%) | 404 (36%) | 279 (40%) | 728 (44%) | 189 (46%) | 848 (42%) | 33 (48%) |  |
| Remote suburbs | 2354 (28%) | 491 (21%) | 174 (15%) | 257 (37%) | 563 (35%) | 147 (36%) | 718 (37%) | 4 (6%) |  |
| Precarious housing | 1366 (16%) | 81 (3%) | 205 (18%) | 32 (5%) | 417 (26%) | 22 (5%) | 593 (30%) | 16 (24%) | <0.001 |

Date expressed in n (%) or median (Q1-Q3); <sup>1</sup>CD4 <350mm<sup>3</sup> or AIDS; <sup>2</sup>CD4 <200mm<sup>3</sup> or AIDS

**Table 2.**
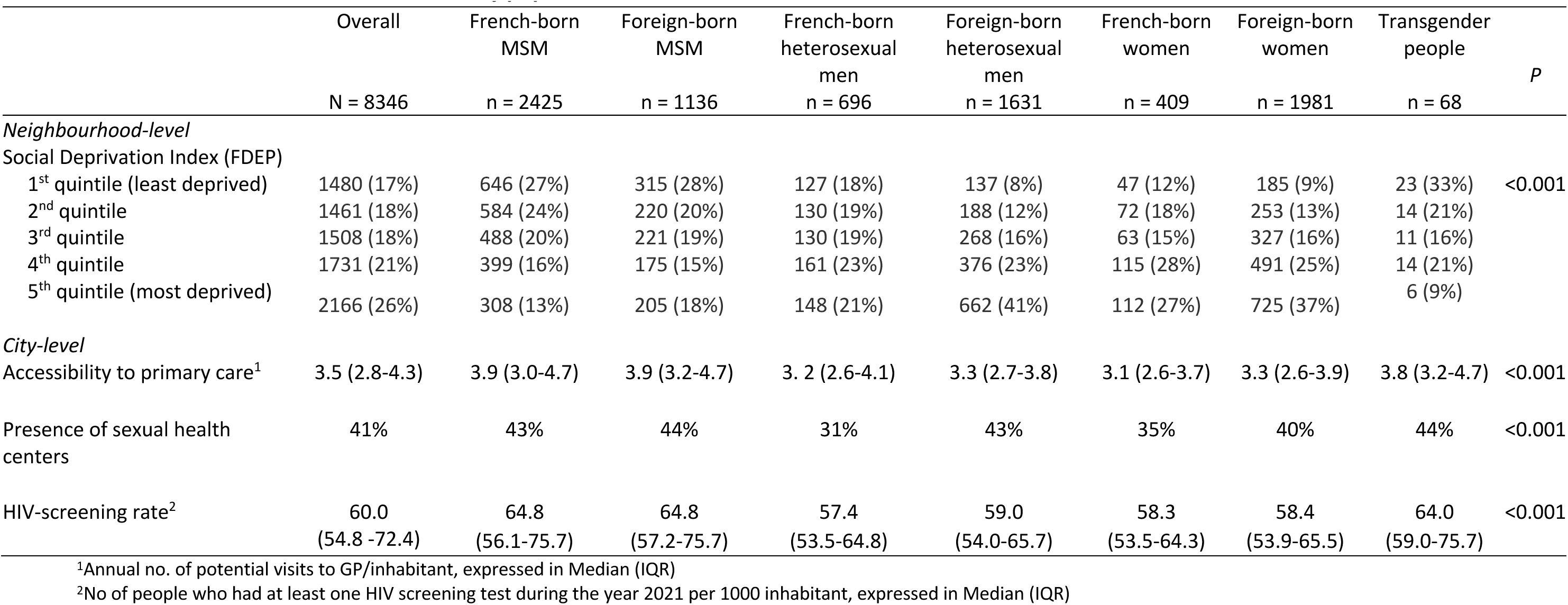
Territorial characteristics of the study population.

LD affected 47% of PLWH, ranging from 30% in FMSM and 40% in fMSM, to 57% in fW and 65% in fHM. Figures 2-3 map new cases and LD prevalence by neighborhood deprivation quintiles, showing LD>45% mainly in the most deprived areas (≥Q4-Q5).

**Figure 2.**
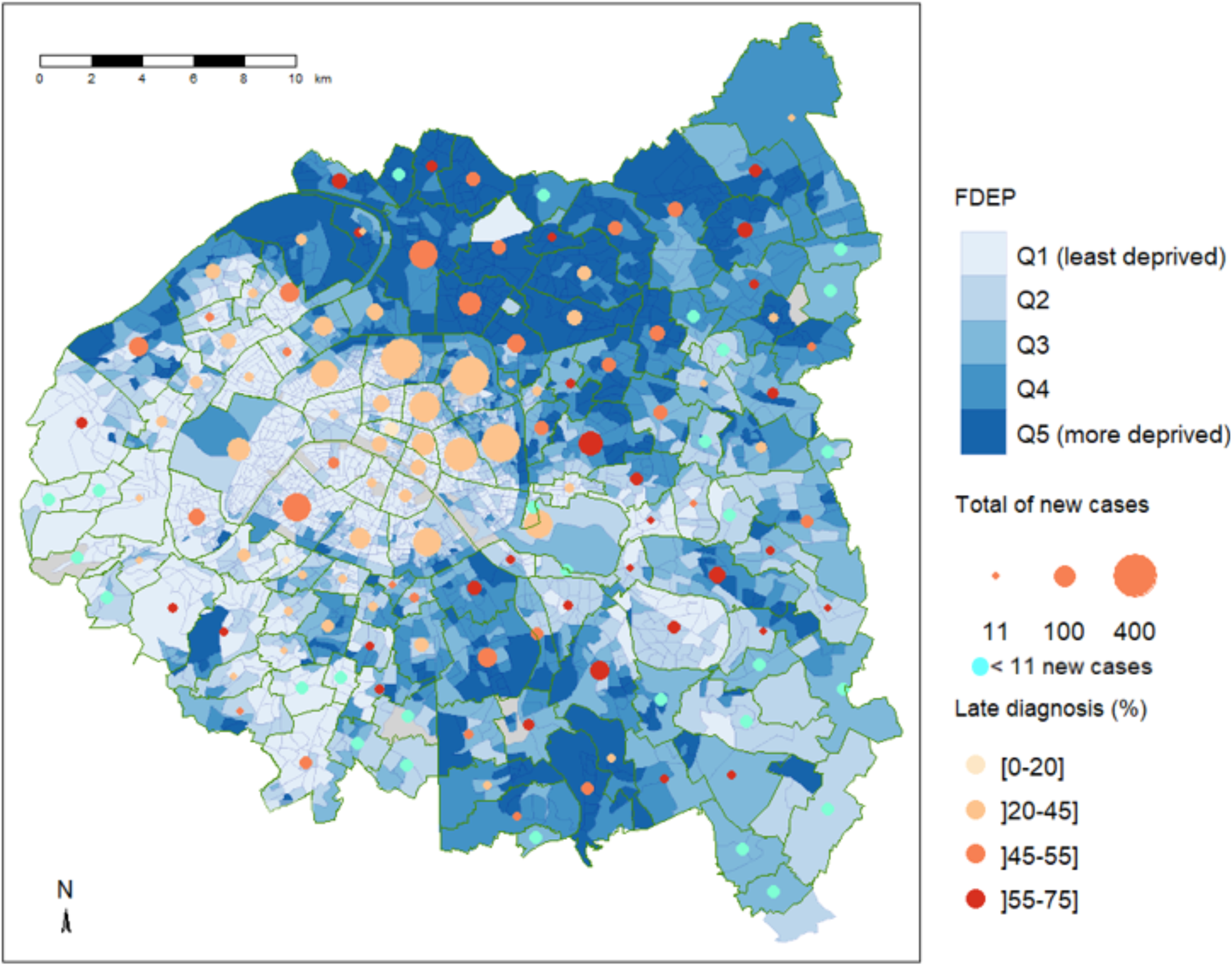
Number of new diagnoses, proportion of late diagnosis and neighborhood deprivation index (FDEP): Inner-Paris and close suburbs

**Figure 3.**
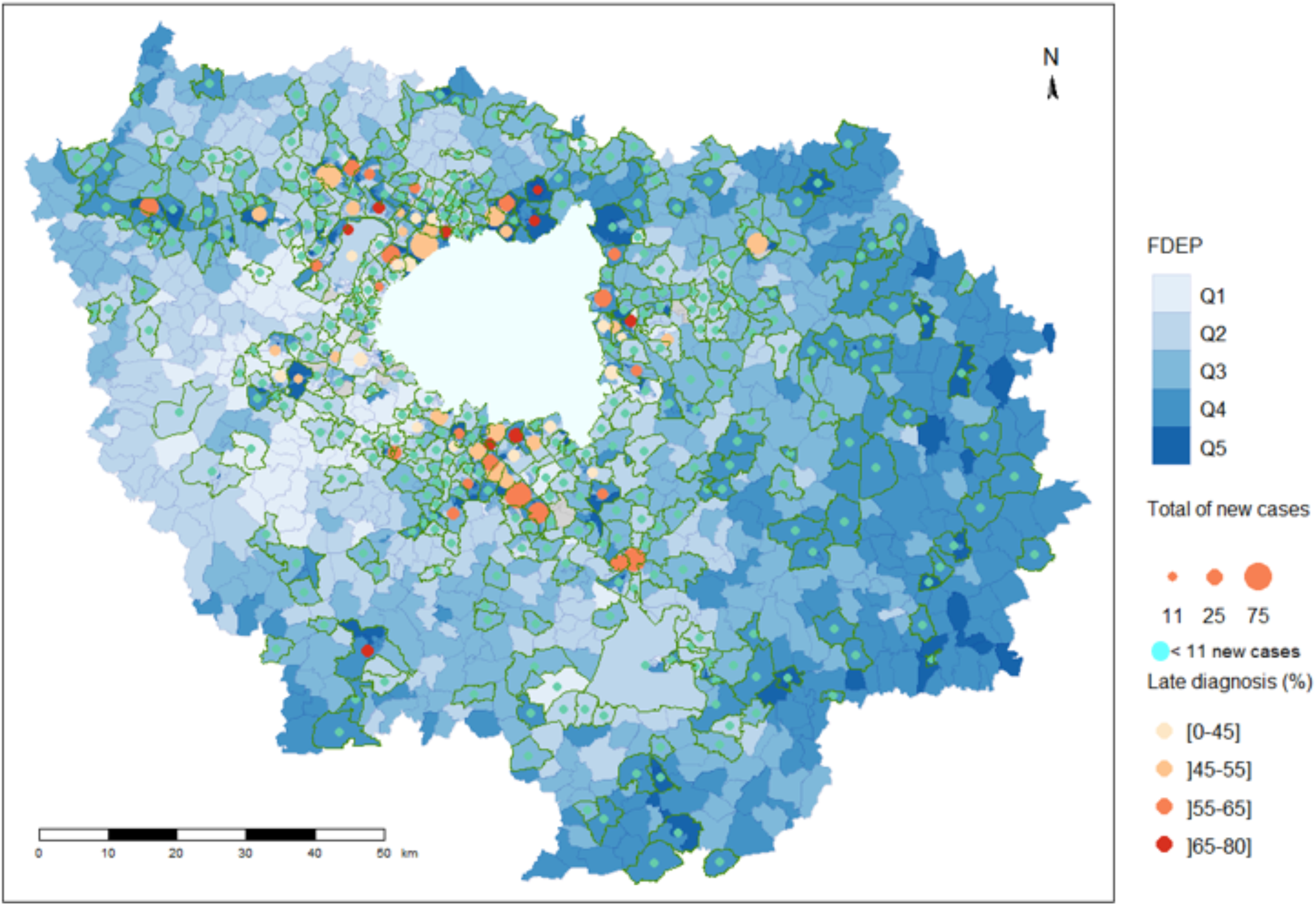
Number of new diagnoses, proportion of late diagnosis and neighborhood deprivation index (FDEP): remote suburbs

### Relationship between FDEP and LD

In univariate analysis (Table 3), all individual and territorial factors were associated with LD. In multilevel multivariate models (Table 4), FDEP remained associated with LD in Models 1-6 (Model 6: aOR_FDEP Q5/Q1_=1.39[1.17–1.65]), independently of primary care access, sexual health center presence, HIV screening rate, residential zone, and precarious housing. After adjusting for transmission group (Model 7), the association was no longer significant. All groups showed higher LD risk than FMSM, especially fHM (aOR=3.67[3.17 - 4.25]) and women (aOR=2.82[2.46-3.24]).

**Table 3.** Factors associated with late HIV diagnosis: univariate analysis (logistic regression)

|  | Late diagnosis<br>n (%) | OR [95% CI] | p |
| --- | --- | --- | --- |
| *Neighbourhood-level* |  |  |  |
| Social Deprivation Index (FDEP) |  |  | <0.001 |
| 1<sup>st</sup> quintile (least deprived) | 581 (39%) | 1 |  |
| 2<sup>nd</sup> quintile | 581 (39%) | 1.02 [0.88- 1.18] |  |
| 3<sup>rd</sup> quintile | 704 (47%) | 1.35 [1.17- 1.57] |  |
| 4<sup>th</sup> quintile | 889 (51%) | 1.63 [1.42- 1.88] |  |
| 5<sup>th</sup> quintile (most deprived) | 1147 (53%) | 1.74 [1.52- 1.99] |  |
| *City-level* |  |  |  |
| Accessibility to primary care<sup>1</sup> |  |  | <0.001 |
| >3.9 – 6.1 | 1192 (40%) | 1 |  |
| ]3.3 – 3.9] | 997 (50%) | 1.47 [1.31-1.65] |  |
| ]2.6 – 3.3] | 860 (49%) | 1.42 [1.26-1.60] |  |
| ≤2.6 | 853 (52%) | 1.61 [1.43-1.82] |  |
| Presence of sexual health centers |  |  | 0.045 |
| No | 2342 (48%) | 1 |  |
| Yes | 1560 (45%) | 0.91 [0.84- 1.00] |  |
| Municipal HIV-screening rate<sup>2</sup> |  |  | <0.001 |
| >64.8 | 1308 (41%) | 1 |  |
| ]58.0 – 64.8] | 945 (50%) | 1.47 [1.31-1.65] |  |
| ]53.5– 58.0] | 867 (50%) | 1.45 [1.29-1.63] |  |
| ≤53.5 | 782 (50%) | 1.47 [1.30-1.66] |  |
| *Individual-level* |  |  |  |
| Residential Zone |  |  | <0.001 |
| Paris inner-city | 1071 (39%) | 1 |  |
| Close suburbs | 1622 (50%) | 1.57 [1.41-1.74] |  |
| Remote suburbs | 1209 (52%) | 1.65 [1.48-1.85] |  |
| Precarious housing |  |  | <0.001 |
| No | 3147 (45%) | 1 |  |
| Yes | 755 (55%) | 1.51 [1.34-1.69] |  |
| Age (per 10 years): median (IQR) | 39 (31-49) | 1.34 [1.29-1.39] | <0.001 |
| Transmission group |  |  | <0.001 |
| French-born MSM | 721 (30%) | 1 |  |
| Foreign-born MSM | 451 (40%) | 1.56 [1.34-1.80] |  |
| French-born heterosexual men | 340 (49%) | 2.26 [1.90-2.68] |  |
| Foreign-born heterosexual men | 1064 (65%) | 4.43 [3.88-5.07] |  |
| French-born women born | 175 (43%) | 1.77 [1.43-2.19] |  |
| Foreign-born women | 1127 (57%) | 3.12 [2.75-3.53] |  |
| Transgender people | 24 (35%) | 1.29 [0.77-2.12] |  |
| Total | 3902 (47%) |  |  |

<sup>1</sup>Annual no. of potential visits to GP/inhabitant, expressed in Median (Q1-Q3)
<sup>2</sup>No of people who had at least one HIV screening test during the year 2021 per 1000 inhabitant, expressed in Median (Q1-Q3)

**Table 4.**
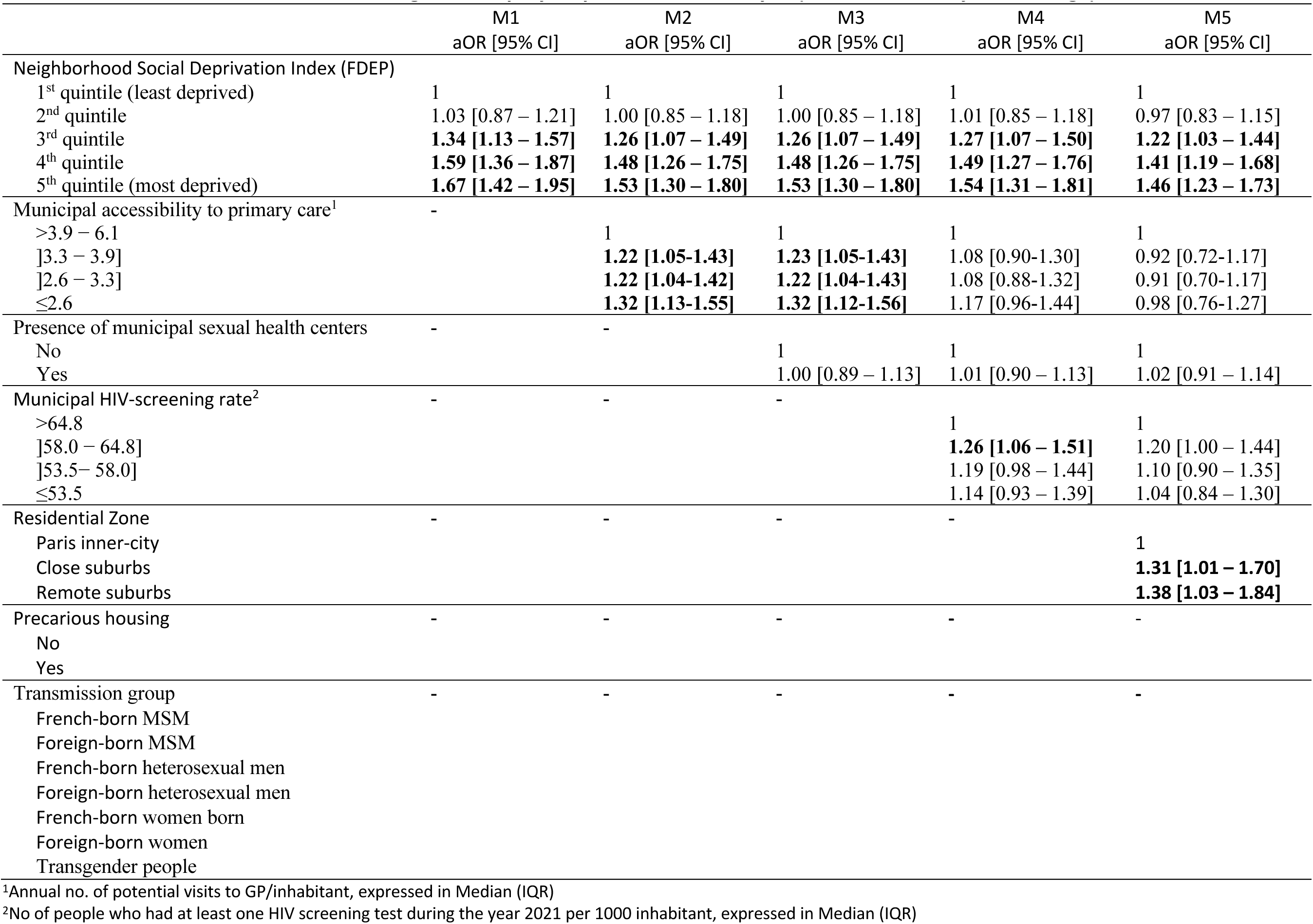
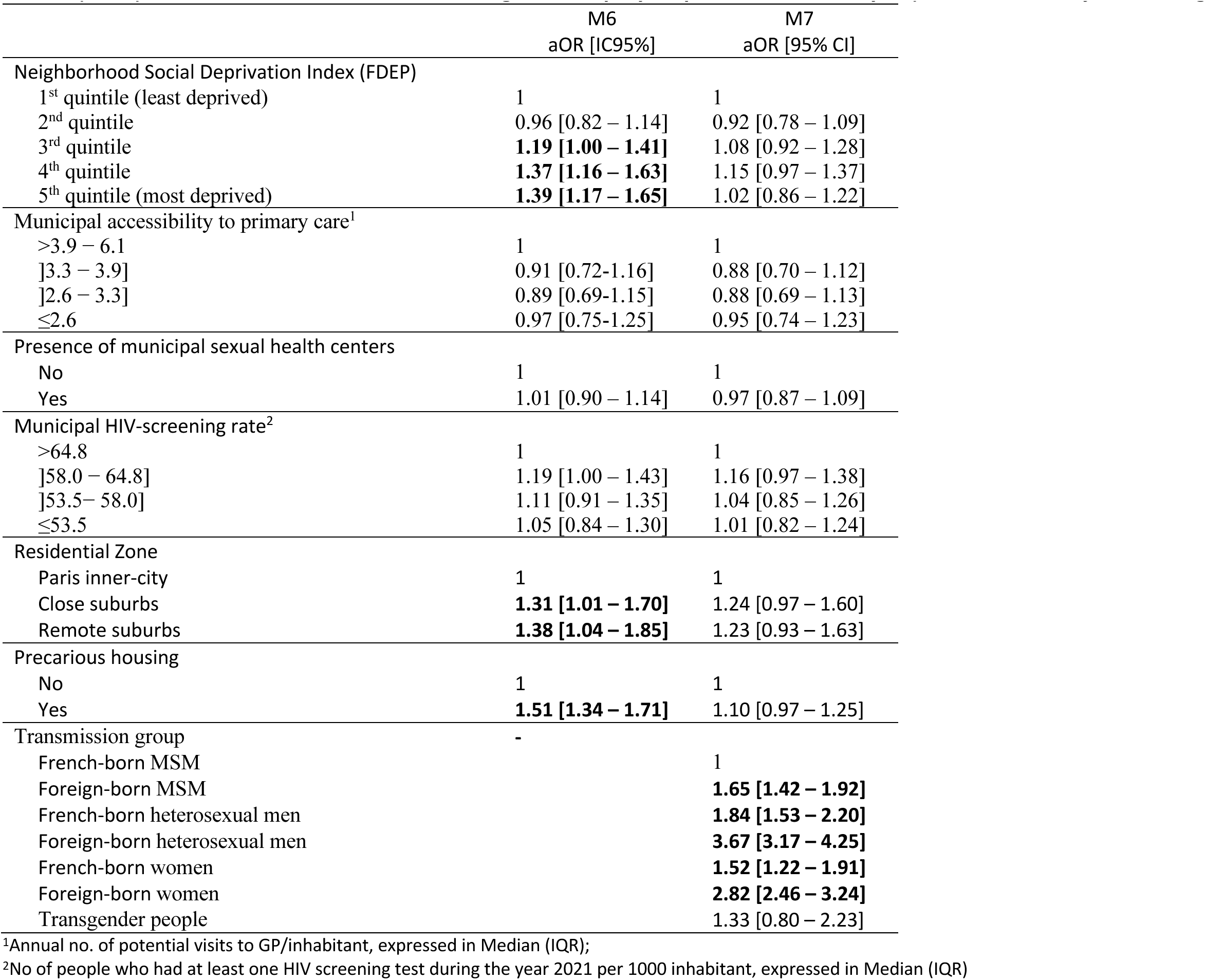
Factors associated with late HIV diagnosis: step-by-step multivariate analysis (all models are adjusted for age)

In the additional analysis (Table 5) including the dichotomized “transmission group” variable, living in deprived neighborhoods remained associated with LD across models.

**Table 5.** Factors associated with late HIV diagnosis: multivariate analysis with transmission risk group categorized as a binary variable (all models being adjusted on age).

|  | M7.1<br>aOR [95% CI] | M7.2<br>aOR [95% CI] | M7.3<br>aOR [95% CI] | M7.4<br>aOR [95% CI] | M7.5<br>aOR [95% CI] | M7.6<br>aOR [95% CI] |
| --- | --- | --- | --- | --- | --- | --- |
| Neighborhood Social Deprivation Index (FDEP) |  |  |  |  |  |  |
| 1<sup>st</sup> quintile (least deprived) | 1 | 1 | 1 | 1 | 1 | 1 |
| 2<sup>nd</sup> quintile | 0.95 [0.81 – 1.12] | 0.96 [0.81 – 1.13] | 0.96 [0.82 – 1.13] | 0.95 [0.80 – 1.12] | 0.97 [0.82 – 1.14] | 0.95 [0.81 – 1.12] |
| 3<sup>rd</sup> quintile | 1.14 [0.97 – 1.35] | **1.19 [1.00 – 1.40]** | **1.19 [1.00 – 1.41]** | 1.14 [0.97 – 1.35] | **1.19 [1.00 – 1.41]** | 1.16 [0.98 – 1.38] |
| 4<sup>th</sup> quintile | **1.24 [1.05 – 1.47]** | **1.36 [1.15 – 1.62]** | **1.37 [1.16 – 1.63]** | **1.30 [1.09 – 1.54]** | **1.38 [1.16 – 1.64]** | **1.32 [1.11 – 1.56]** |
| 5<sup>th</sup> quintile (most deprived) | **1.18 [1.00 – 1.40]** | **1.38 [1.16 – 1.63]** | **1.39 [1.17 – 1.64]** | **1.24 [1.04 – 1.47]** | **1.39 [1.17 – 1.65]** | **1.31 [1.10 – 1.55]** |
| Municipal accessibility to primary care<sup>1</sup> |  |  |  |  |  |  |
| >3.9 – 6.1 | 1 | 1 | 1 | 1 | 1 | 1 |
| ]3.3 – 3.9] | 0.89 [0.70 – 1.12] | 0.91 [0.72 – 1.16] | 0.91 [0.72 – 1.15] | 0.89 [0.71 – 1.13] | 0.91 [0.72 – 1.15] | 0.91 [0.72 – 1.16] |
| ]2.6 – 3.3] | 0.89 [0.69 – 1.14] | 0.89 [0.69 – 1.15] | 0.89 [0.69 – 1.15] | 0.87 [0.68 – 1.12] | 0.89 [0.69 – 1.15] | 0.90 [0.70 – 1.16] |
| ≤2.6 | 0.95 [0.74 – 1.23] | 0.97 [0.75 – 1.25] | 0.97 [0.75 – 1.25] | 0.96 [0.74 – 1.23] | 0.97 [0.75 – 1.26] | 0.97 [0.75 – 1.25] |
| Presence of municipal sexual health centers |  |  |  |  |  |  |
| No | 1 | 1 | 1 | 1 | 1 | 1 |
| Yes | 1.00 [0.89 – 1.12] | 1.01 [0.90 – 1.14] | 1.01 [0.90 – 1.14] | 0.99 [0.88 – 1.11] | 1.01 [0.90 – 1.14] | 1.01 [0.90 – 1.13] |
| Municipal HIV-screening rate<sup>2</sup> |  |  |  |  |  |  |
| >64.8 | 1 | 1 | 1 | 1 | 1 | 1 |
| ]58.0 – 64.8] | 1.15 [0.96 – 1.37] | 1.19 [1.00 – 1.43] | 1.20 [1.00 – 1.43] | 1.21 [1.01 – 1.45] | 1.20 [1.00 – 1.44] | 1.18 [0.98 – 1.41] |
| ]53.5 – 58.0] | 1.04 [0.86 – 1.27] | 1.11 [0.91 – 1.35] | 1.11 [0.91 – 1.35] | 1.12 [0.92 – 1.37] | 1.11 [0.91 – 1.35] | 1.07 [0.88 – 1.31] |
| ≤53.5 | 0.99 [0.81 – 1.23] | 1.05 [0.85 – 1.30] | 1.05 [0.85 – 1.30] | 1.07 [0.86 – 1.32] | 1.05 [0.85 – 1.30] | 1.03 [0.83 – 1.27] |
| Residential Zone |  |  |  |  |  |  |
| Paris inner-city | 1 | 1 | 1 | 1 | 1 | 1 |
| Close suburbs | 1.27 [0.99 – 1.64] | **1.30 [1.01 – 1.69]** | **1.31 [1.01 – 1.70]** | 1.28 [0.99 – 1.66] | **1.32 [1.02 – 1.71]** | **1.30 [1.01 – 1.68]** |
| Remote suburbs | 1.31 [0.99 – 1.74] | **1.37 [1.03 – 1.83]** | **1.39 [1.04 – 1.85]** | **1.33 [1.00 – 1.77]** | **1.39 [1.04 – 1.86]** | **1.35 [1.01 – 1.79]** |
| Precarious housing |  |  |  |  |  |  |
| No | 1 | 1 | 1 | 1 | 1 | 1 |
| Yes | **1.24 [1.09 – 1.40]** | **1.51 [1.34 – 1.71]** | **1.51 [1.33 – 1.70]** | **1.40 [1.23 – 1.58]** | **1.49 [1.32 – 1.69]** | **1.39 [1.22 – 1.57]** |
| Transmission group |  |  |  |  |  |  |
| Ref (Others) | 1 | 1 | 1 | 1 | 1 | 1 |
| Transmission group (one by one) | **0.43 [0.39 – 0.48]** | 0.90 [0.79 – 1.03] | 0.96 [0.82 – 1.14] | **2.08 [1.84 – 2.34]** | **0.77 [0.62 – 0.95]** | **1.52 [1.36 – 1.70]** |
|  | French-born MSM | Foreign-born MSM | French-born heterosexual men | Foreign-born heterosexual men | French-born women | Foreign-born women |

<sup>1</sup>Annual no. of potential visits to GP/inhabitant, expressed in Median (Q1-Q3)
<sup>2</sup>No of people who had at least one HIV screening test during the year 2021 per 1000 inhabitant, expressed in Median (Q1-Q3)

## Discussion

This study confirmed the high prevalence of late diagnosis (LD) among PLWH newly diagnosed in Greater Paris (2014-2021) and highlights marked socio-territorial inequalities and disparities by gender, transmission mode, and country of birth.

LD prevalence was 47%, consistent with recent European estimates (2023): 53% in the EU/EEA^24^, 47% in Western Europe^24^, 43% in France and 44-48% in Greater Paris^21,25^. Such high rates reflect excessive delay between contamination and diagnosis, pointing to insufficient and/or inadequate HIV screening and prevention. LD remained frequent among MSM (30% among FMSM, 40% among fMSM), who represent 43% of new diagnoses in Greater Paris, and was even higher among foreign-born heterosexuals (65% among men, 57% among women), who accounted for an additional 43% of new cases. In France, heterosexual men and migrants are at higher risk of LD and the situation has not improved over time^26^. This echoes a 2010 representative population-based survey in Greater Paris showing 43% of men and 33% of women had never been tested for HIV^27^, particularly in disadvantaged and immigrant-dense neighborhoods. Interestingly, people of sub-Saharan origin were most likely to have been recently tested^28^. This aligns with recent French data showing a short median delay (five months in 2023) between arrival in France and diagnosis among immigrant with HIV^29^, suggesting that LD stems from a lack of screening before or during migration. However, for the 51% of foreign-born PLWH contaminated in France, LD likely reflects insufficient local testing^29,30^. Community-based and outreach testing has proven effective at reaching people who had never been tested previously, with high acceptability of rapid testing when conducted outside traditional healthcare settings^31,32,33^. Developing outreach-based screening strategies is probably essential.

We subsequently observed that almost half of PLWH resided in the most deprived neighborhood, compared with 39% of the general population in Greater Paris. This was associated with a 40% higher LD risk, independently of primary care access, sexual health centers, screening rates, residential zone, and housing conditions. These territorial inequalities call for innovative, geography-based, strategies by targeting the most deprived areas and key populations. The ANRS-MIE COINCIDE cohort enabled the development of an online interactive map of new HIV diagnoses in Greater Paris (available since 2024)^21^, This tool supports, geography-based strategies by identifying priority areas for screening and prevention — those most affected and most socioeconomically deprived according to our findings (Figures 2-3). Our results contrast with a previous French study reporting no association between neighborhood deprivation and LD^15^. However, that analysis used national deprivation quintiles, with 70% of participants living in the two most deprived quintiles, likely masking the deprivation effect on LD. In our study, we used quintiles derived from the study population itself, which likely better captured local disparities.

Moreover, the “transmission group” variable as defined by our seven categories classification, integrating gender, region of birth, and transmission mode, emerged as a relevant social indicator. Most European studies considered only gender and transmission mode, leaving aside region of birth despite strong interrelations^34,35,36^. After inclusion of the transmission group in our final model, the FDEP-LD association disappeared, suggesting confounding by population composition: groups with higher LD risk are concentrated in more deprived areas. MSM and foreign-born non-MSM each represented 43% of new diagnoses but lived in very different settings: half of MSM in inner Paris versus 79% of foreign-born non-MSM in the suburbs. Also, while one third of MSM lived in the most deprived neighborhoods, this proportion doubled among non-MSM groups. Thus, territorial LD disparities largely reflect the unequal geographic distribution of social groups.

Finally, when using the dichotomized transmission group, the FDEP-LD association persisted across models, confirming that deprivation independently contributes to LD risk, and that the seven-category classification provides a more meaningful social indicator unlike the dichotomized variable. Future studies on health inequalities among PLWH should consider such composite categorizations depending on objectives.

This study has some limitations. First, individual social data are not available in medical records, which would nevertheless allow to distinguish contextual effects from those related to specific social PLWH characteristics^37,38,39,40^. Furthermore, our results may not be generalizable beyond Greater Paris, even if it is reasonable to assume that the disparities observed in this study could also be found in other urban areas with a high level of social diversity. Strengths include comprehensive inclusion and geocoding of all new HIV cases across 61 centers between 2014 and 2021, providing an almost exhaustive regional dataset. Another strength is the use of the “transmission group” variable in the models, which allows the formation of socially and epidemiologically homogeneous groups and enables a more detailed characterization of at-risk subpopulations.

In conclusion, socioeconomic inequalities continue to drive the HIV epidemic in Greater Paris, with higher LD prevalence in deprived areas. Despite extensive free screening policies, higher LD rates among foreign-born non-MSM PLWH highlights that current strategies are failing to sufficiently reach these key populations. Innovative strategies combining population-based and geography-driven approaches are needed. Intensifying outreach-based screening and prevention in foreign key populations, by targeting the most affected and deprived areas, is undoubtedly central.

## Ethics

This study was approved in 2019 by the Ethics Evaluation Committee of Inserm and by the Committee of Experts for Research, Studies, and Evaluations in the Field of Health (CESREES).

## Acknowledgements

We thank Valérie Ferron, Adrien Saunal, and Laetitia Firdion from the Observatoire Régional de Santé-Ile-de-France for their valuable contributions in providing contextual data for this study. We are grateful to all ANRS-MIE COINCIDE study participants and research assistants, without whom this work would not have been possible.

